## Supplementary files for "Characteristics and six-month viral load suppression of clients presenting with advanced HIV disease in South Africa"

**Supplementary Table 1. Socio-demographic characteristics of PREFER study participants with or without baseline CD4 count**

| **Characteristic** | **Level** | **Patients without baseline CD4 count** | **Clients with baseline CD4 count** | **Total** |
| --- | --- | --- | --- | --- |
| N (%) |  | n= 160 (15) | n = 938 (85) | (N = 1098) |
| Median age (IQR) |  | 35 (29-42) | 33 (27-40) | 33 (27-41) |
| Age | 18-24years | 19 (12) | 169 (18) | 188 (17) |
|  | 25-49 years | 124 (78) | 677 (72) | 801 (73) |
|  | 50+ years | 17 (11) | 92 (10) | 109 (10) |
| Sex | Male | 51 (32) | 261 (28) | 312 (28) |
|  | Female | 109 (68) | 677 (72) | 786 (72) |
| Marital status | Married living with partner | 58 (36) | 289 (31) | 347 (32) |
|  | Married but not living with partner | 64 (40) | 441 (47) | 505 (46) |
|  | Single | 38 (24) | 208 (22) | 246 (22) |
| Education | Primary or less | 62 (39) | 347 (37) | 409 (37) |
|  | Secondary | 72 (45) | 453 (48) | 525 (48) |
|  | Post-secondary | 26 (16) | 138 (15) | 164 (15) |
| Occupation | Formal | 36 (22) | 204 (22) | 240 (22) |
|  | Informal | 29 (18) | 187 (20) | 216 (20) |
|  | Unemployed | 85 (53) | 477 (51) | 562 (51) |
|  | Student/Trainee | 10 (6) | 70 (7) | 80 (7) |
| Food scarcity | Never | 114 (71) | 658 (70) | 772 (70) |
|  | Seldom | 13 (8) | 59 (6) | 72 (7) |
|  | Sometimes | 29 (18) | 191 (20) | 220 (20) |
|  | Often | 4 (2) | 30 (3) | 34 (3) |
| Access to money for health care | Very difficult | 24 (15) | 127 (14) | 151 (14) |
|  | Difficult | 72 (45) | 392 (42) | 464 (42) |
|  | Easy | 57 (36) | 378 (40) | 435 (40) |
|  | Very easy | 7 (4) | 41 (4) | 48 (4) |
| Facility patient volume | <2000 TROA | 55 (34) | 236 (25) | 291 (27) |
|  | 2000-4000 TROA | 56 (35) | 371 (40) | 427 (39) |
|  | >4000 TROA | 49 (31) | 331 (35) | 380 (35) |
| Patient ART history | Initiating today | 38 (24) | 273 (29) | 311 (28) |
|  | Re-engagement | 33 (21) | 135 (14) | 168 (15) |
|  | On treatment | 89 (56) | 530 (57) | 619 (56) |

**Supplementary Table 2. Socio-demographic characteristics of HIV clients stratified by six months viral load documentation**

| **Characteristic** | **Level** | | **Patients without six-month viral load** | | **Clients with six-month viral load** | | **Total** | |
| --- | --- | --- | --- | --- | --- | --- | --- | --- |
| N (%) |  | | n = 314(33) | | n= 624 (67) | | N = 938 | |
| Median age (IQR) | |  | | 35 (29-42) | | 33 (27-40) | | 33 (27-41) |
| Age group | | 18-24 years | | 63 (20) | | 106 (17) | | 169 (18) |
|  | | 25-49 years | | 230 (73) | | 447 (72) | | 677 (72) |
|  | | 50+ years | | 21 (7) | | 71 (11) | | 92 (10) |
| Sex | | Male | | 97 (31) | | 164 (26) | | 261 (28) |
|  | | Female | | 217 (69) | | 460 (74) | | 677 (72) |
| Marital status | | Married living with partner | | 94 (30) | | 195 (31) | | 289 (31) |
|  | | Married but not living with partner | | 137 (44) | | 304 (49) | | 441 (47) |
|  | | Single | | 83 (26) | | 125 (20) | | 208 (22) |
| Education | | Primary or less | | 127 (40) | | 220 (35) | | 347 (37) |
|  | | Secondary | | 137 (44) | | 316 (51) | | 453 (48) |
|  | | Post-secondary | | 50 (16) | | 88 (14) | | 138 (15) |
| Occupation | | Formal | | 62 (20) | | 142 (23) | | 204 (22) |
|  | | Informal | | 62 (20) | | 125 (20) | | 187 (20) |
|  | | Unemployed | | 169 (54) | | 308 (49) | | 477 (51) |
|  | | Student/Trainee | | 21 (7) | | 49 (8) | | 70 (7) |
| Food scarcity | | Never | | 211 (67) | | 447 (72) | | 658 (70) |
|  | | Seldom | | 23 (7) | | 36 (6) | | 59 (6) |
|  | | Sometimes | | 73 (23) | | 118 (19) | | 191 (20) |
|  | | Often | | 7 (2) | | 23 (4) | | 30 (3) |
| Access to money for health care | | Very difficult | | 39 (12) | | 88 (14) | | 127 (14) |
|  | | Difficult | | 149 (47) | | 243 (39) | | 392 (42) |
|  | | Easy | | 115 (37) | | 263 (42) | | 378 (40) |
|  | | Very easy | | 11 (4) | | 30 (5) | | 41 (4) |
| Facility patient volume | | <2000 TROA | | 65 (21) | | 171 (27) | | 236 (25) |
|  | | 2000-4000 TROA | | 118 (38) | | 253 (41) | | 371 (40) |
|  | | >4000 TROA | | 131 (42) | | 200 (32) | | 331 (35) |
| Patient ART history | | Initiating today | | 124 (39) | | 149 (24) | | 273 (29) |
|  | | Re-engagement | | 46 (15) | | 89 (14) | | 135 (14) |
|  | | On treatment | | 144 (46) | | 386 (62) | | 530 (57) |
